# Geographic patterns of the number of root canals in permanent molars. A Systematic Review

**DOI:** 10.1101/2024.06.08.24308647

**Authors:** Alain Manuel Chaple Gil, Meylin Santiesteban Velázquez, Kelvin Ian Afrashtehfar

## Abstract

**Introduction:** The variability in the distribution and number of root canals in human permanent molars makes endodontic therapy difficult if the prevalence of these patterns is not well known.

**Objective:** To synthesize scientific evidence on geographical variations in the number of root canals in permanent molars.

**Methods:** A study conducted at the University of Medical Sciences of Havana analyzed the number of root canals in permanent molars. We searched for primary studies published in English, Spanish, or Portuguese that reported the number of canals in each tooth. We excluded clinical case studies, editorials, and studies that did not provide specific information on duct anatomy. A sensitivity analysis was performed to assess the impact of the methodological quality of the studies in the two groups. The findings were synthesized by grouping the studies by geographical region and describing the prevalence of different root canal configurations.

**Results:** Data on permanent molars were collected from 93 studies that contributed 60402 molars studied. In Africa, the first and second upper molars commonly have three root canals (45.62% and 41.31%, respectively), similar to the first and second lower molars (59.79% and 67.55%, respectively). In America, the upper first molar most frequently has four root canals (56.66%). In Asia, it is more common to find that all molar groups have three canals (51.09%– 65.58%), except for the lower thirds, which may have two or three canals (42.88% and 57.12%). In Europe, all molars are frequently present with three canals, except for the lower third. In Oceania, the first upper molars commonly have six root canals (65%).

**Conclusion:** Significant variations in the number of root canals in permanent molars according to geographic region were observed. Distinct patterns were observed in the Americas, Asia, Africa, Europe, and Oceania, with varying prevalences of different root canal configurations.

## INTRODUCTION

The controversy regarding the morphology of the roots and root canals of teeth poses significant challenges for endodontic practice, as a comprehensive judgment of this branch of dental science is required to obtain successful treatment results. Therefore, several classification systems have been described to accurately characterize root and root canal configurations^(1, 2, 3)^; however, this is not sufficient. So much so that Studies that have used advanced imaging techniques, such as micro-computed tomography (micro-CT) and cone beam computed tomography (CBCT), have revealed the complexities of root canal anatomy, highlighting the limitations of the existing classification systems^(4, 5)^. In addition, investigations into the morphology of the root and canal of the third molars have revealed new canal configurations not previously described, emphasizing the unpredictable nature of tooth anatomy and the need for further research in this field.^(4, 6)^

The number of root canals in permanent molars varies depending on the type of tooth and the individual anatomy. Research indicates that maxillary and mandibular molar primaries can have a wide range of root and channel configurations, with the most common being three canals and three roots in maxillary first molars.^(7)^ and two separate roots with two distinct mesial canals and a distal canal for the first mandibular ^(8)^ Other studies conclude that a significant proportion of the first permanent maxillary molars have three roots and four canals, with a high prevalence of a second mesiobuccal canal (MB2)^(9, 10, 11)^ That’s why understanding these variations is vitally important to achieving successful endodontic treatments.

Most universities advocate that their curricula have an internationalization that provides knowledge about their graduates’ ways of acting to be able to perform in any context that they have to live in their professional lives.^(12, 13)^ That is why the present research pays tribute to this globalization of knowledge in the dental guild, since a dentist graduated in America could practice his profession in patients from the Middle East and know the frequency of the number of root canals that the molars will have to whichtheye will perform endodonticsatn that latitude of the planet.

The variability in the distribution and number of root canals in human permanent molars makes endodontic therapy difficult if the prevalence of these patterns is not well known. For this reason, making available to the dental community a vision of how these patterns behave by geographical area would contribute to the dental sciences.

As a result of the previous approaches, the problem to be dealt with in this research is outlined in the scientific question: Are there significant differences in the morphology and configuration of root canals of permanent molars between different geographic populations?

To answer this question, the objective of this research was to synthesize the available scientific evidence on the geographical variations in the internal anatomy of the root canals of permanent molars in terms of their number, establishing significant patterns, and differences between different populations.

## METHODS

A systematic review study was conducted from April to July 2024 at the University of Medical Sciences of Havana, where this project was approved by the Scientist Committee. The PRISMA guide was used for the execution of the research, and the databases used for the retrieval of the reports were PubMed, Scopus, and Web of Science (WOS).

### Inclusion criteria

Primary research studies were included (originals, investigations, research papers or clinical trials) These had to show reports of anatomical aspects related to the number of root canals of the permanent molars. Articles in English, Spanish, and Portuguese were also included. Articles that did not specifically report the number of canals per permanent molar but could be determined by inference were also included.

### Exclusion Criteria

Articles on clinical cases, opinion articles, editorials, reviews, or others that differed from those described in the inclusion criteria were excluded. Studies that deal with the subject studied primary or deciduous teeth, studies that do not report anatomical information on the number of root canals, and other articles written in languages other than English, Spanish, or Portuguese.

In addition, were excluded articles that were not available in full text and that did not show at least in the abstract relevant results for the extraction of data necessary in the research.

### Search for studies

The formulations used in PubMed was: ((Pulp Canal*[Title]) OR (Root Canal*[Title]) OR (dental canal*[Title]) OR (canal*[Title])) AND ((anatomy [Title]) OR (morphology [Title])), and with the same keywords, it was used in the databases consulted, adjusting them to the characteristics of each one.

Alternatively, the Research Rabbit website (https://researchrabbitapp.com/home) was used to retrieve articles that were not the result of the searches but were relevant to the study. The phrase used was ‘root canal morphology of human molars.’

### Procedures

Calibration was performed among the authors to evaluate the selected articles. The degree of coincidence of the evaluations made by the reviewers was determined using *Orwin’s* method in 1994, and a kappa statistic was performed to measure the agreement among the reviewers who would make simple decisions about inclusion/exclusion. Kappa values between 0.40 and 0.59 were considered to reflect acceptable agreement, 0.60 to 0.74 to be an adequate agreement, and 0.75 or more to reflect excellent agreement.

The records obtained from the databases were processed into a document from the bibliographic manager EndNote, through which export files were managed for the screening process on the Rayyan® site (https://rayyan.ai/). Using this tool, duplicates and other records that did not match the primary inclusion/exclusion criteria were eliminated, and the researchers blindly determined which articles were suitable for inclusion and which were not. In case of discrepancy or doubt, a 3rd author was in charge of deriving the inclusion results.

### Data Extraction

From the included reports, data such as the author and year of publication, design of the study, age of the study population, geographic location of the study, number of molars analyzed, dental group (1st, 2nd, and 3rd upper and/or lower permanent molars), and number of molar groups with one, two, three, four, five, six, or more root canals were obtained.

Sensitivity analysis was performed to assess the impact of the methodological quality of the studies on the results, which were divided into two groups: 1) high-quality group, which included studies that used high-resolution imaging techniques (CT or CBCT) to evaluate root anatomy; and 2) low-quality group, which included studies that relied on conventional radiographs or less accurate assessment methods.

### Data analysis

This systematic review focuses on the quantitative synthesis of the results of primary studies not to estimate a particular effect, but to generalize about the number of root canals in permanent molars globally. As the main objective was to provide a summary estimate of the outcome, it was not necessary to conduct a detailed risk of bias analysis (ROB) of each individual study. Despite not including a formal ROB, the following steps were taken to ensure the quality of the included evidence: rigorous study selection, critical evaluation of the studies, analysis of heterogeneity, and sensitivity of analysis.

A narrative synthesis of the findings was made, grouping the studies by geographic regions and describing the reported prevalence of different root canal configurations (number of canals) in maxillary and mandibular molars for each population.

Global and regional heat maps, plots, and tables were generated, showing the geographic distribution of the prevalence of different root canal configurations in maxillary and mandibular molars. This allows the spatial patterns to be clearly visualized.

For the sensitivity analysis of the studies, we performed descriptive statistics separately for each group and compared the results to assess the impact of the methodological quality.

RStudio® 2024.04.1 Build 748 was used for data processing. The data frames and RStudio codes used for the processing of information are available at the following link according to the principles of open science (https://doi.org/10.17632/xvgf59n75k.1):

All the results were grouped into tables, maps, and graphs showing relative and absolute frequencies for a better understanding of the patterns of the number of canals for each molar group in each geographical region.

## RESULTS

Calibration of the authors who reviewed the articles included in the study was excellent in all cases.

The search yielded 2917 articles distributed among PubMed (1086), Scopus (1240), and WOS (559). Only 32 articles were obtained from the Research Rabbit. (Figure 1)

**Figure 1.**
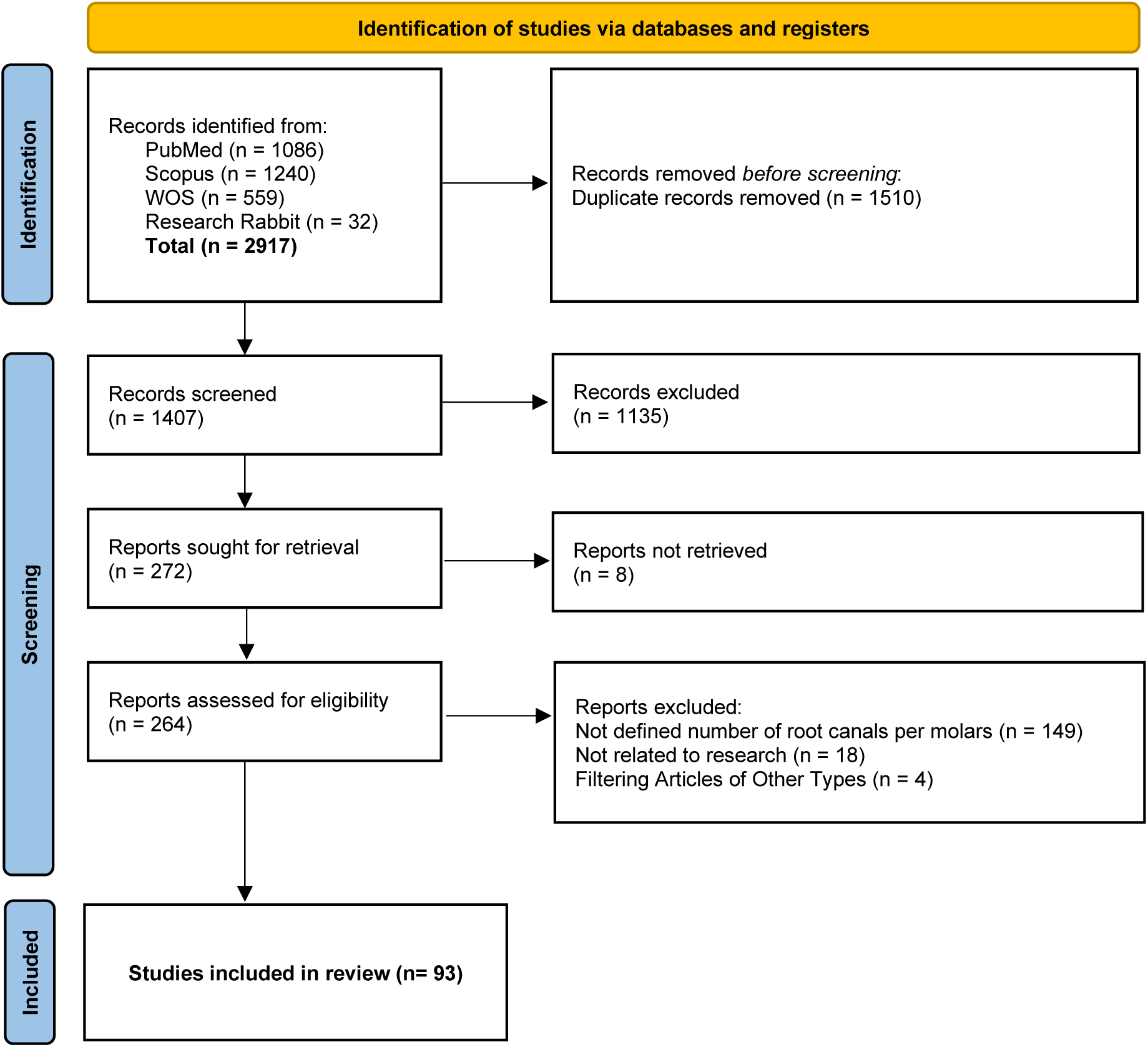
PRISMA flow diagram to select reports to include in the systematic review

After eliminating duplicates and decanting articles that did not meet the inclusion criteria, 272 reports were obtained that could be evaluated for eligibility. Of the 149 articles, the number of root canals per tooth could not be defined, 8 were not available in full text, 18 were not related to the research, and 4 were articles of other modalities reflected in the exclusion criteria (Figure 1). At the end of the screening, 93 reports were obtained that were included in this study and are listed in Table 1.

**Table 1.**
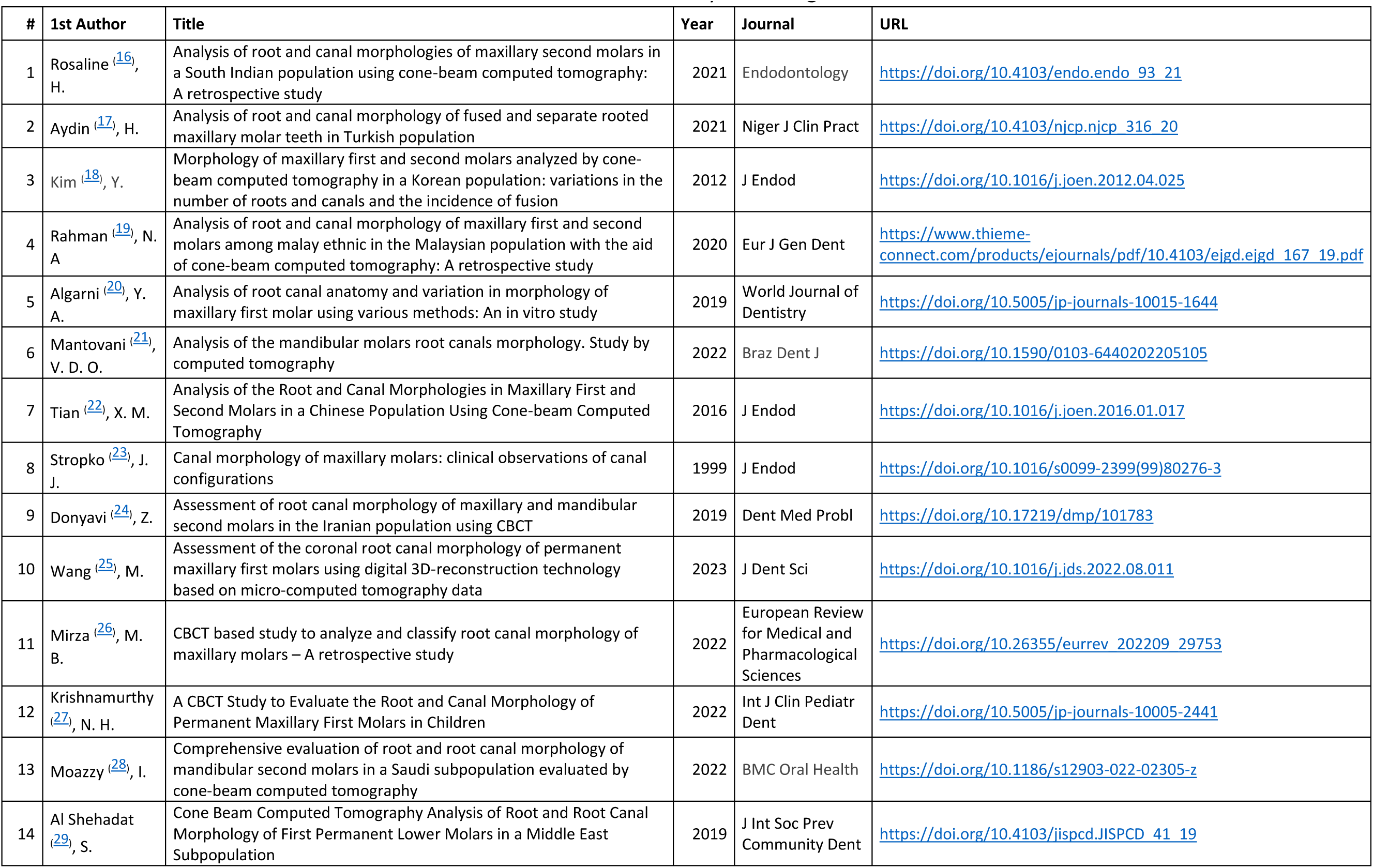
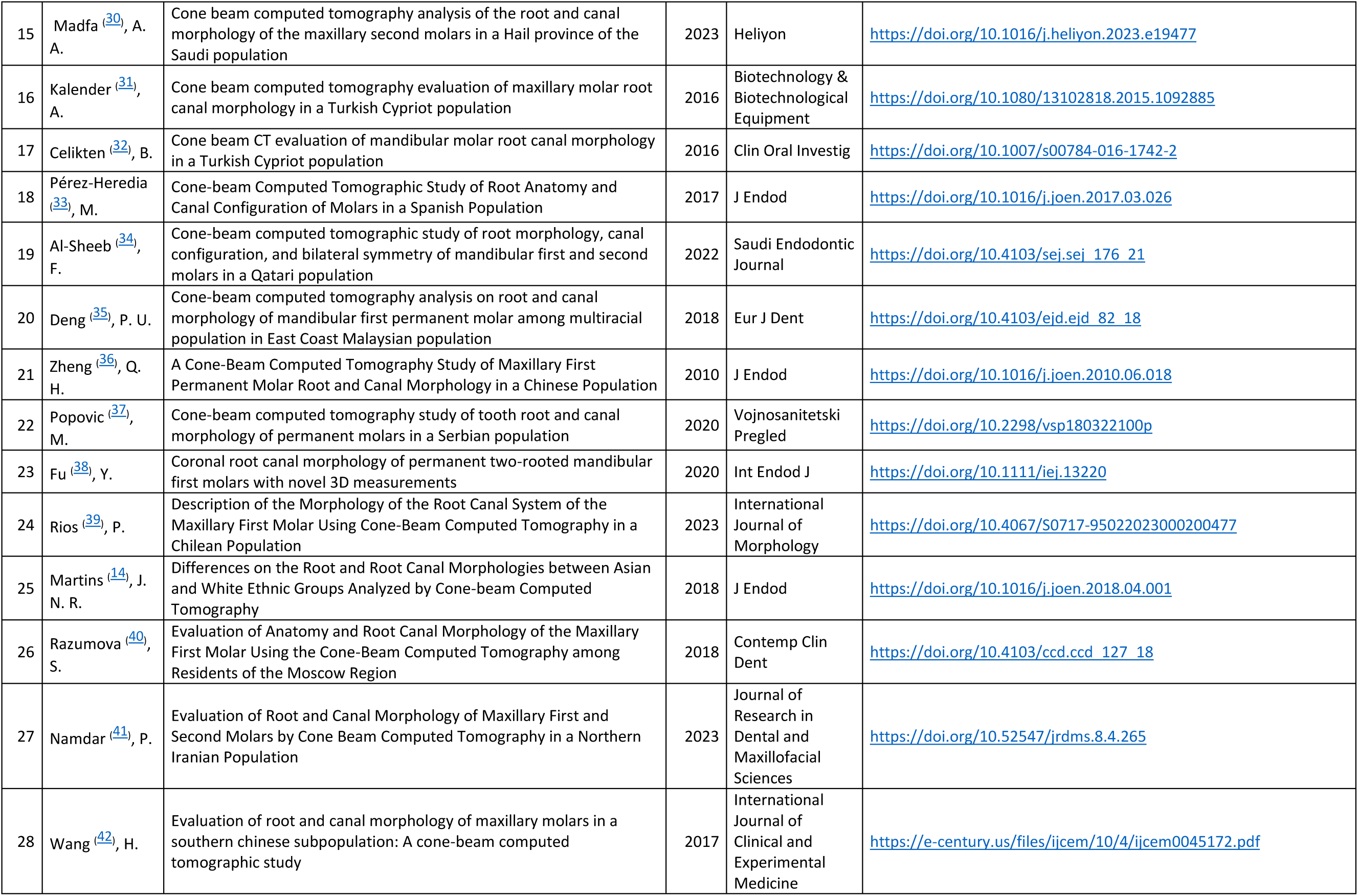
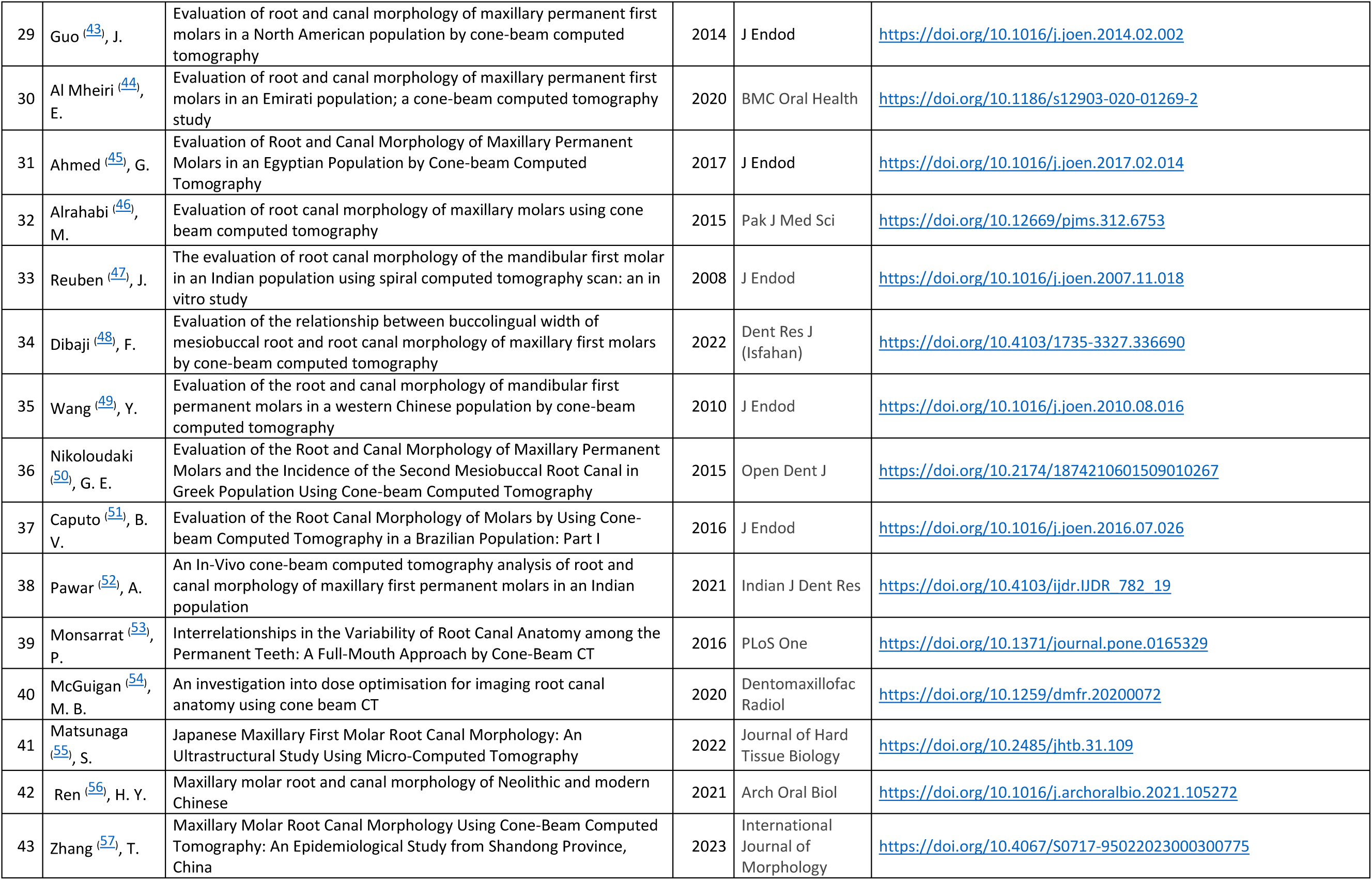
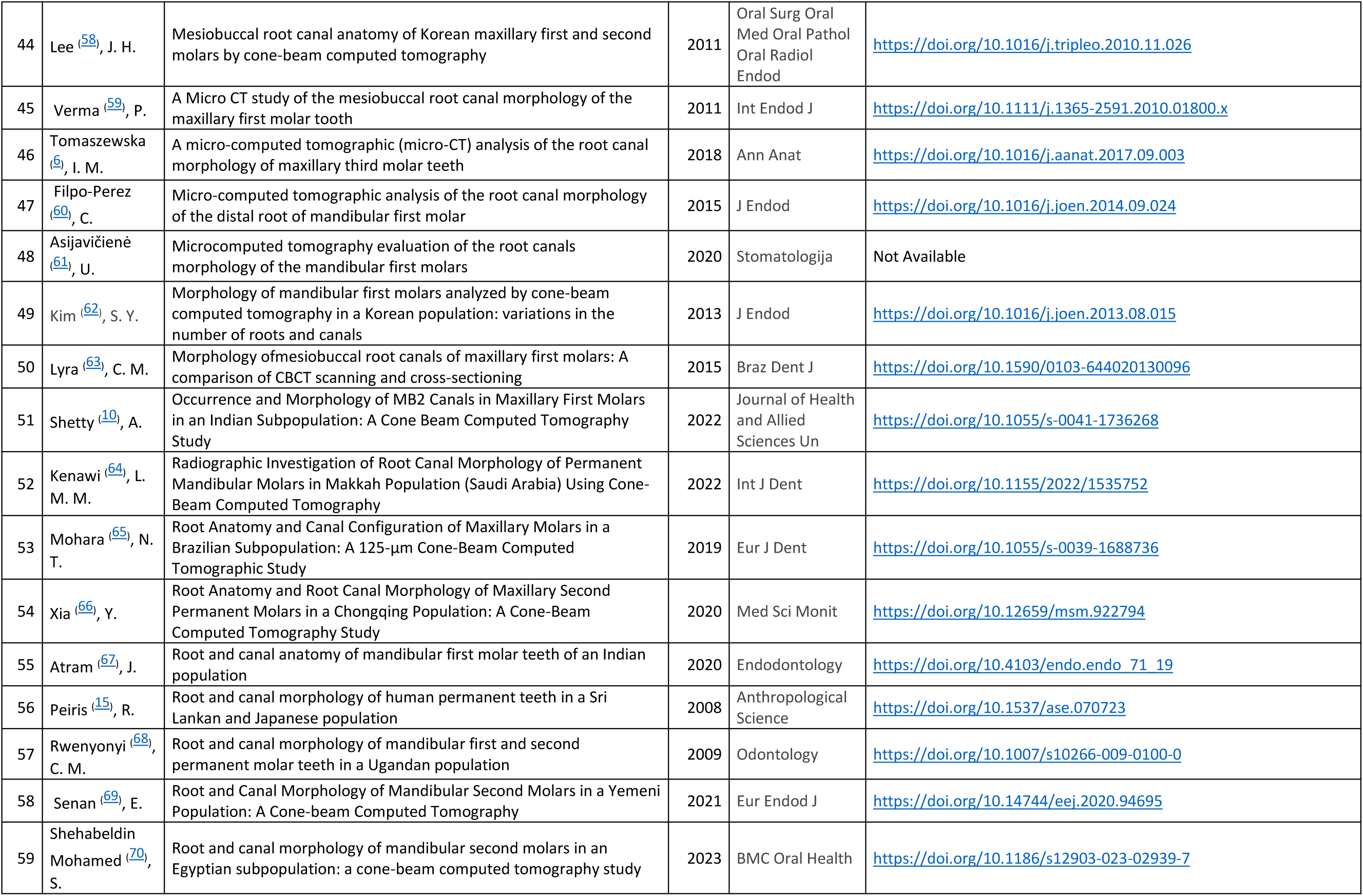
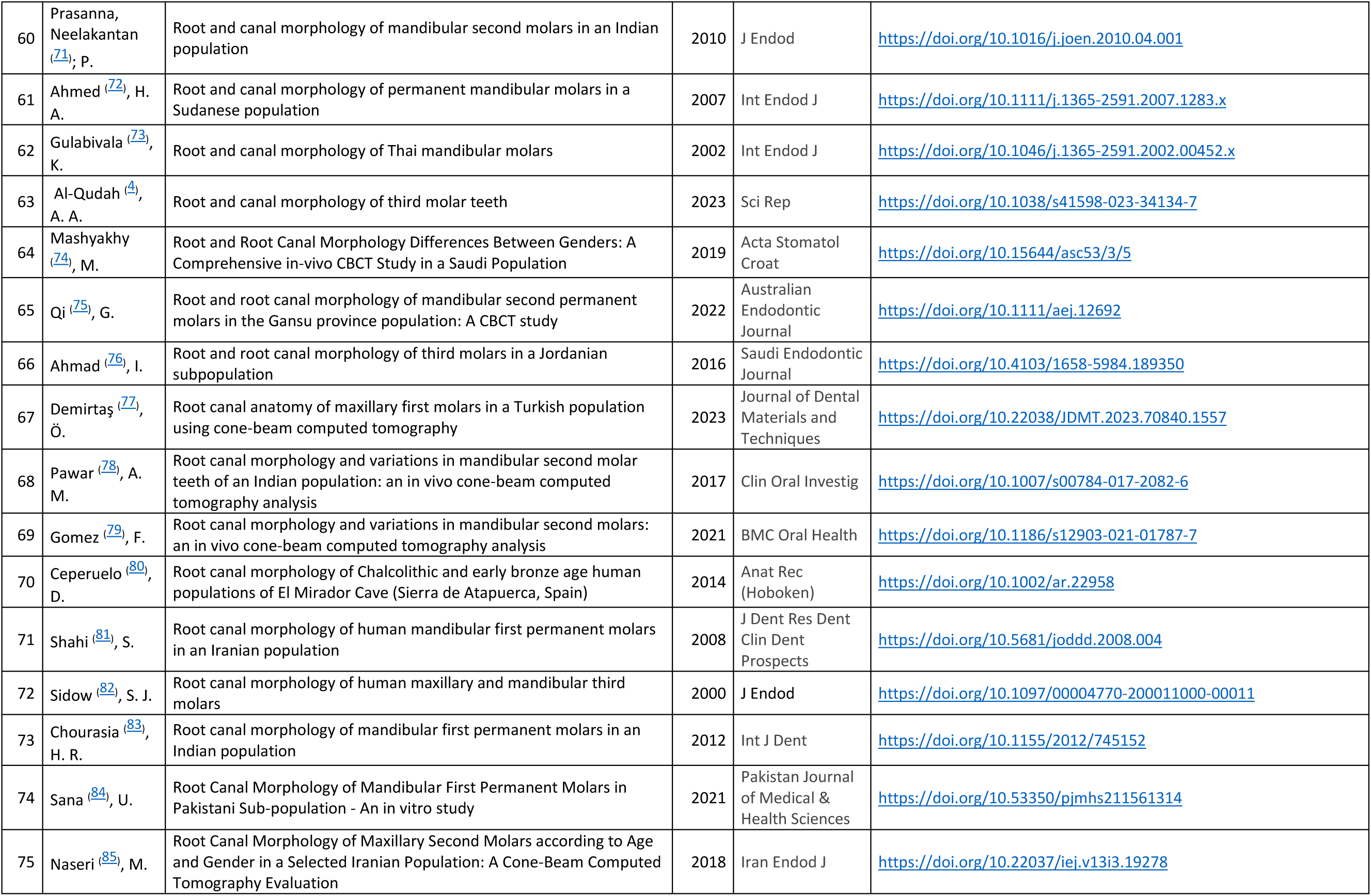
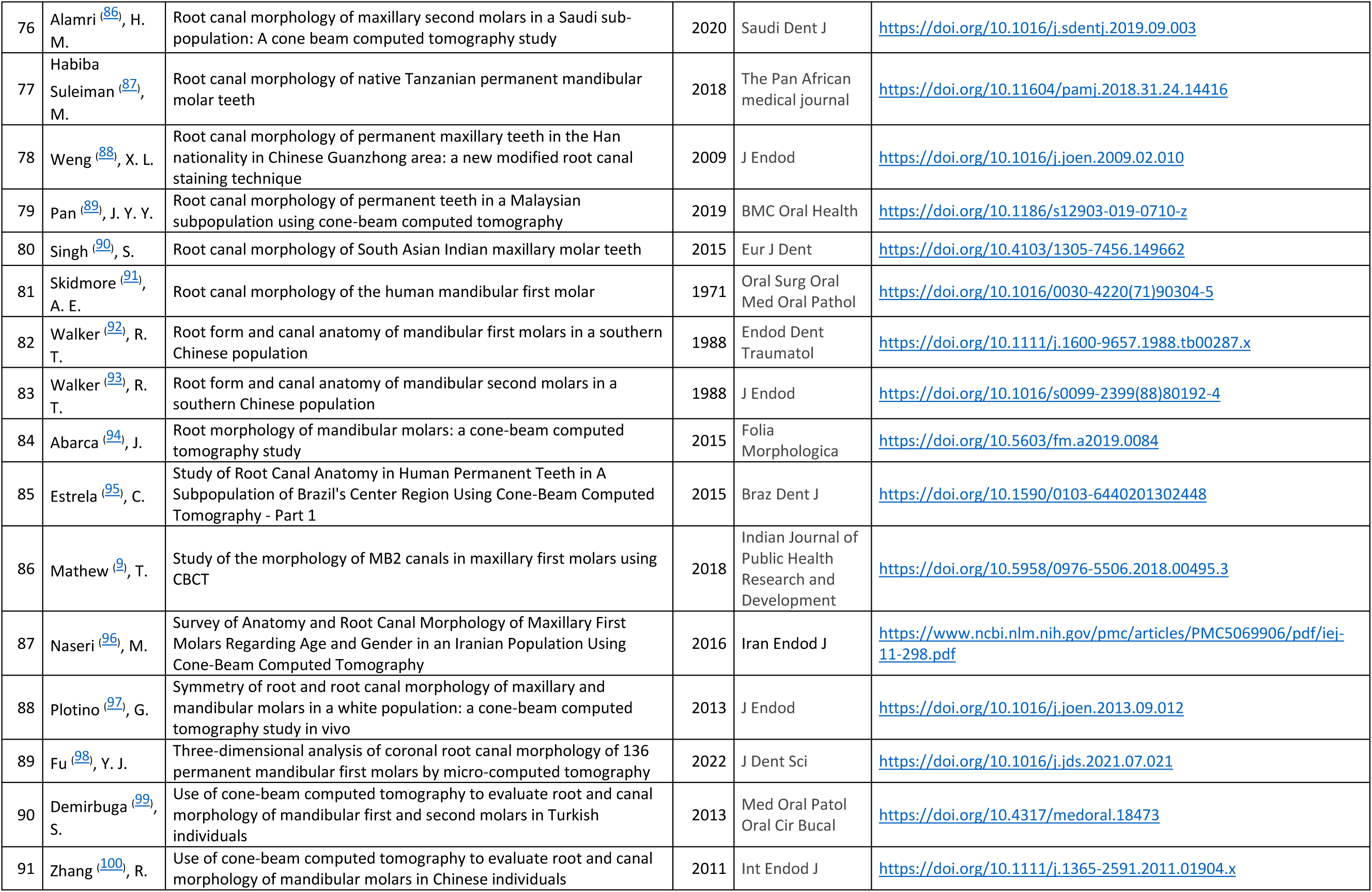
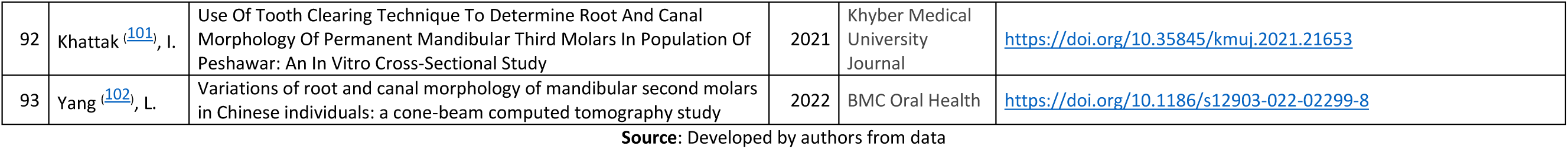
Distribution of the articles included in the study according to relevant data for their identification.

These articles yielded information on 95 sample records of molars from countries in which the number of canals per tooth could be determined. This was due to the fact that the studies of Martins et al^(14)^ and Peiris et al^(15)^ studied two populations from different countries.

Only one study was written in Spanish, while the rest were published in English. The Journal of Endodontics was the most frequent journal (19.35%), where reports were published, and there were no significant frequencies of authors. (Table 1)

Sensitivity analysis of the studies was not necessary because only 10.75% of the studies belonged to the group with low measurement quality.

Articles from 1971 to 2023 were included, with 2022 being the year with the highest number of articles published (12 articles), followed by 2020 (9 articles), 2021, and 2023 with 8 articles each. (Table1 & Figure 2)

**Figure 2.**
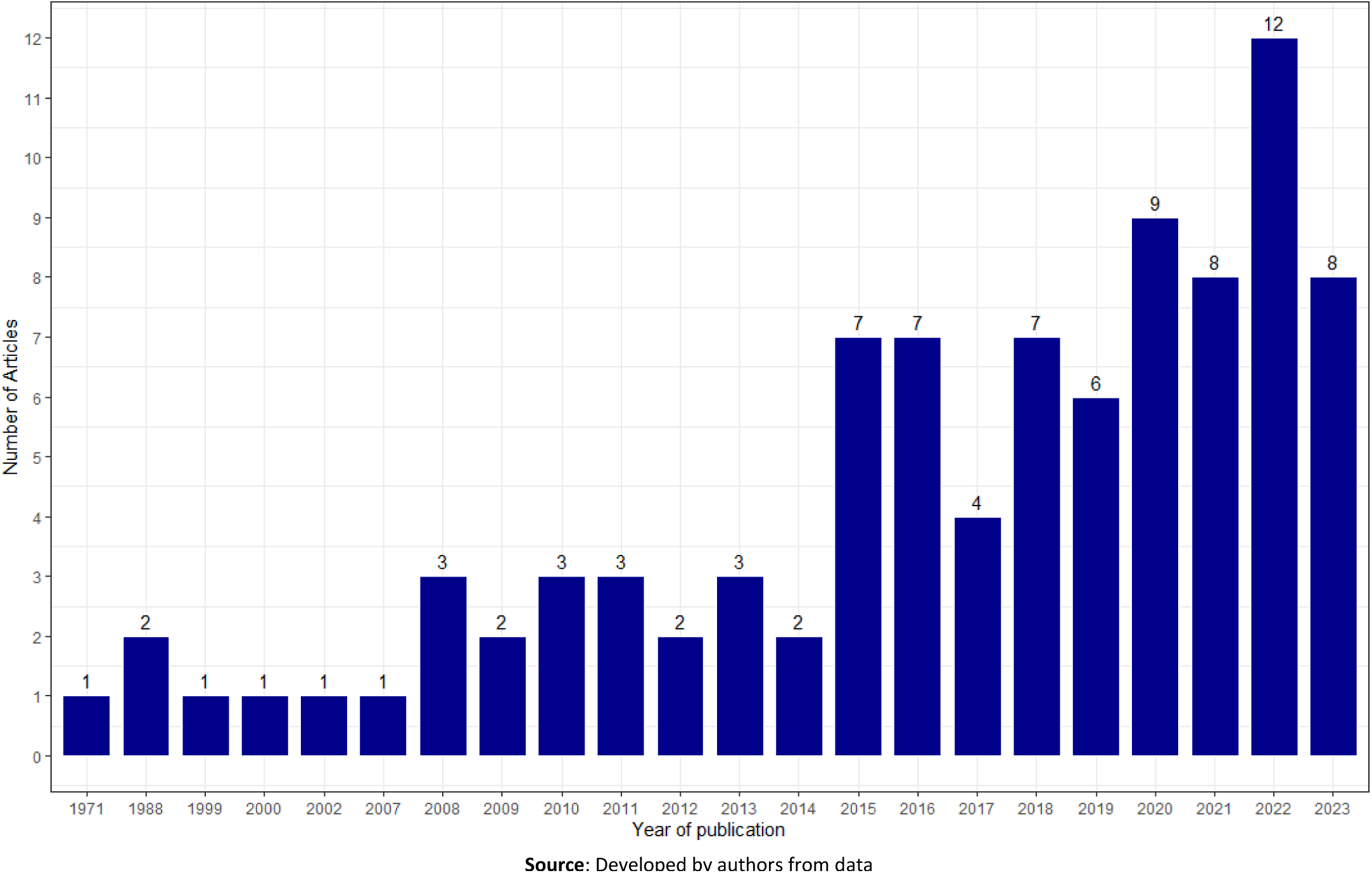
Distribution of number of articles published per year.

Data were extracted from 60402 molars from all dental groups (Figure 3). China had the highest number of molars, representing 23.88% of the total. This was followed by Saudi Arabia, South Korea, India, and Iran, with 7.94%, 7.37%, 7.21%, and 7.03%, respectively. According to the data extracted, Ireland had the lowest number of molars contributed, with three molars, followed by Poland with 0.13%, Lithuania 0.10%, and New Zealand with 0.03% (Table 2).

**Figure 3.**
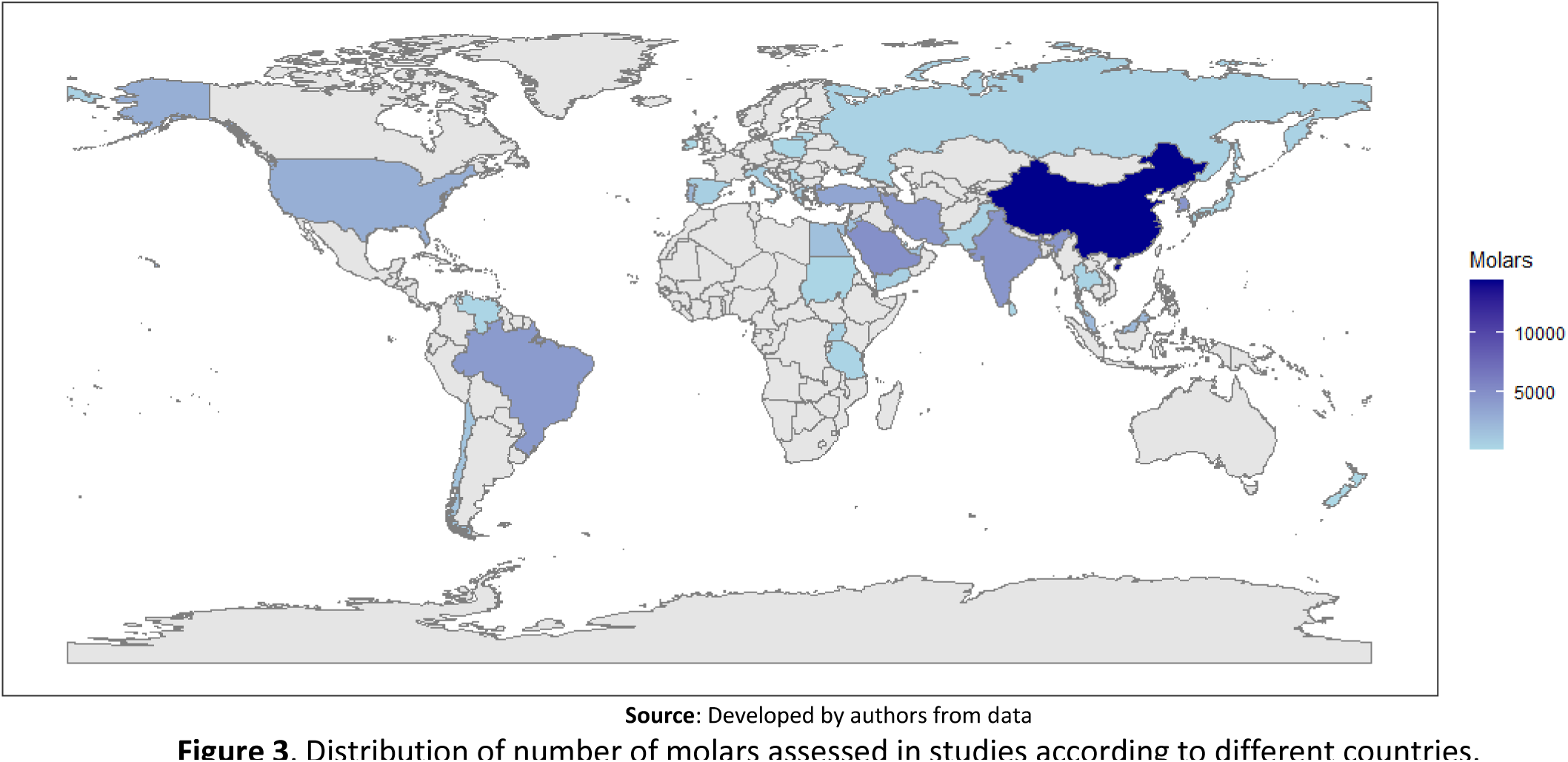
Distribution of number of molars assessed in studies according to different countries.

**Table 2.** Distribution of molars by arch and countries involved.

| Countries<br>(ISO2) | Total of Molars |  | 1<sup>st</sup> Upper Molar |  | 2<sup>nd</sup> Upper Molar |  | 3<sup>th</sup> Upper Molar |  | 1<sup>st</sup> Lower Molar |  | 2<sup>nd</sup> Lower Molar |  | 3<sup>th</sup> Lower Molar |  |
| --- | --- | --- | --- | --- | --- | --- | --- | --- | --- | --- | --- | --- | --- | --- |
|  | # | % | # | % | # | % | # | % | # | % | # | % | # | % |
| AE | 1329 | 2,20 | 522 | 2,60 | 0 | - | 0 | - | 807 | 7,64 | 0 | - | 0 | - |
| BR | 4053 | 6,71 | 1143 | 5,70 | 1038 | 6,01 | 0 | - | 1157 | 10,95 | 715 | 6,98 | 0 | - |
| CL | 1221 | 2,02 | 199 | 0,99 | 0 | - | 0 | - | 510 | 4,83 | 512 | 5,00 | 0 | - |
| CN | 14423 | 23,88 | 6054 | 30,21 | 5180 | 30,01 | 43 | 4,02 | 1203 | 11,39 | 1943 | 18,96 | 0 | - |
| CY | 1616 | 2,68 | 373 | 1,86 | 438 | 2,54 | 0 | - | 384 | 3,64 | 421 | 4,11 | 0 | - |
| EG | 1565 | 2,59 | 605 | 3,02 | 610 | 3,53 | 0 | - | 0 | - | 350 | 3,42 | 0 | - |
| ES | 580 | 0,96 | 155 | 0,77 | 150 | 0,87 | 0 | - | 138 | 1,31 | 137 | 1,34 | 0 | - |
| FR | 589 | 0,98 | 149 | 0,74 | 167 | 0,97 | 0 | - | 130 | 1,23 | 143 | 1,40 | 0 | - |
| GR | 812 | 1,34 | 410 | 2,05 | 402 | 2,33 | 0 | - | 0 | - | 0 | - | 0 | - |
| HK | 200 | 0,33 | 100 | 0,50 | 100 | 0,58 | 0 | - | 0 | - | 0 | - | 0 | - |
| IE | 3 | 0,00 | 3 | 0,01 | 0 | - | 0 | - |  | - | 0 | - | 0 | - |
| IN | 4352 | 7,21 | 1692 | 8,44 | 600 | 3,48 | 100 | 9,35 | 761 | 7,21 | 1199 | 11,70 | 0 | - |
| IR | 4246 | 7,03 | 836 | 4,17 | 2339 | 13,55 | 0 | - | 209 | 1,98 | 862 | 8,41 | 0 | - |
| IT | 596 | 0,99 | 161 | 0,80 | 157 | 0,91 | 0 | - | 117 | 1,11 | 161 | 1,57 | 0 | - |
| JO | 1390 | 2,30 | 0 | - | 0 | - | 681 | 63,64 | 0 | - | 0 | - | 709 | 58,16 |
| JP | 153 | 0,25 | 100 | 0,50 | 53 | 0,31 | 0 | - | 0 | - | 0 | - | 0 | - |
| KR | 4452 | 7,37 | 1271 | 6,34 | 1242 | 7,19 | 0 | - | 1939 | 18,36 | 0 | - | 0 | - |
| LK | 213 | 0,35 | 0 | - | 213 | 1,23 | 0 | - | 0 | - | 0 | - | 0 | - |
| LT | 62 | 0,10 | 0 | - | 0 | - | 0 | - | 62 | 0,59 | 0 | - | 0 | - |
| MY | 2015 | 3,34 | 824 | 4,11 | 890 | 5,16 | 0 | - | 301 | 2,85 | 0 | - | 0 | - |
| NZ | 20 | 0,03 | 20 | 0,10 | 0 | - | 0 | - | 0 | - | 0 | - | 0 | - |
| PK | 393 | 0,65 | 0 | - | 0 | - | 0 | - | 200 | 1,89 | 0 | - | 193 | 15,83 |
| PL | 78 | 0,13 | 0 | - | 0 | - | 78 | 7,29 | 0 | - | 0 | - | 0 | - |
| PT | 2242 | 3,71 | 714 | 3,56 | 589 | 3,41 | 0 | - | 463 | 4,38 | 476 | 4,65 | 0 | - |
| QA | 450 | 0,75 | 0 | - | 0 | - | 0 | - | 195 | 1,85 | 255 | 2,49 | 0 | - |
| RS | 138 | 0,23 | 138 | 0,69 | 0 | - | 0 | - | 0 | - | 0 | - | 0 | - |
| RU | 410 | 0,68 | 410 | 2,05 | 0 | - | 0 | - | 0 | - | 0 | - | 0 | - |
| SA | 4793 | 7,94 | 1447 | 7,22 | 1781 | 10,32 | 0 | - | 575 | 5,44 | 990 | 9,66 | 0 | - |
| SD | 200 | 0,33 | 0 | - | 0 | - | 0 | - | 100 | 0,95 | 100 | 0,98 | 0 | - |
| TH | 351 | 0,58 | 0 | - | 0 | - | 0 | - | 118 | 1,12 | 60 | 0,59 | 173 | 14,19 |
| TR | 3391 | 5,61 | 940 | 4,69 | 703 | 4,07 | 0 | - | 823 | 7,79 | 925 | 9,03 | 0 | - |
| TZ | 231 | 0,38 | 0 | - | 0 | - | 0 | - | 146 | 1,38 | 85 | 0,83 | 0 | - |
| UG | 447 | 0,74 | 0 | - | 0 | - | 0 | - | 224 | 2,12 | 223 | 2,18 | 0 | - |
| US | 2698 | 4,47 | 1775 | 8,86 | 611 | 3,54 | 168 | 15,70 | 0 | - | 0 | - | 144 | 11,81 |
| VE | 190 | 0,31 | 0 | - | 0 | - | 0 | - | 0 | - | 190 | 1,85 | 0 | - |
| YE | 500 | 0,83 | 0 | - | 0 | - | 0 | - | 0 | - | 500 | 4,88 | 0 | - |
| **Total** | **60402** | **100,00** | **20041** | **33,18** | **17263** | **28,58** | **1070** | **1,77** | **10562** | **17,49** | **10247** | **16,96** | **1219** | **2,02** |

AE (United Arab Emirates) BR (Brazil) CL (Chile) CN (China) CY (Cyprus) EG (Egypt) ES (Spain) FR (France) GR (Greece) HK (Hong Kong) IE (Ireland) IN (India) IR (Iran) IT (Italy) JO (Jordan) JP (Japan) KR (South Korea) LK (Sri Lanka) LT (Lithuania) MY (Malaysia) NZ (New Zealand) PK (Pakistan) PL (Poland) PT (Portugal) QA (Qatar) RS (Serbia) RU (Russia) SA (Saudi Arabia) SD (Sudan) TH (Thailand) TR (Turkey) TZ (Tanzania, United Republic of) UG (Uganda) US (United States of America) VE (Venezuela) YE (Yemen)
**Source:** Developed by authors from data
Source: Developed by authors from data

The most represented dental group of molars was the upper first molars with 33.18% of the total, and the ones with the lowest reported by countries were the upper third molars (1.77 %) and lower molars (2.02 %) (Table 2 and Figure 4).

**Figure 4.**
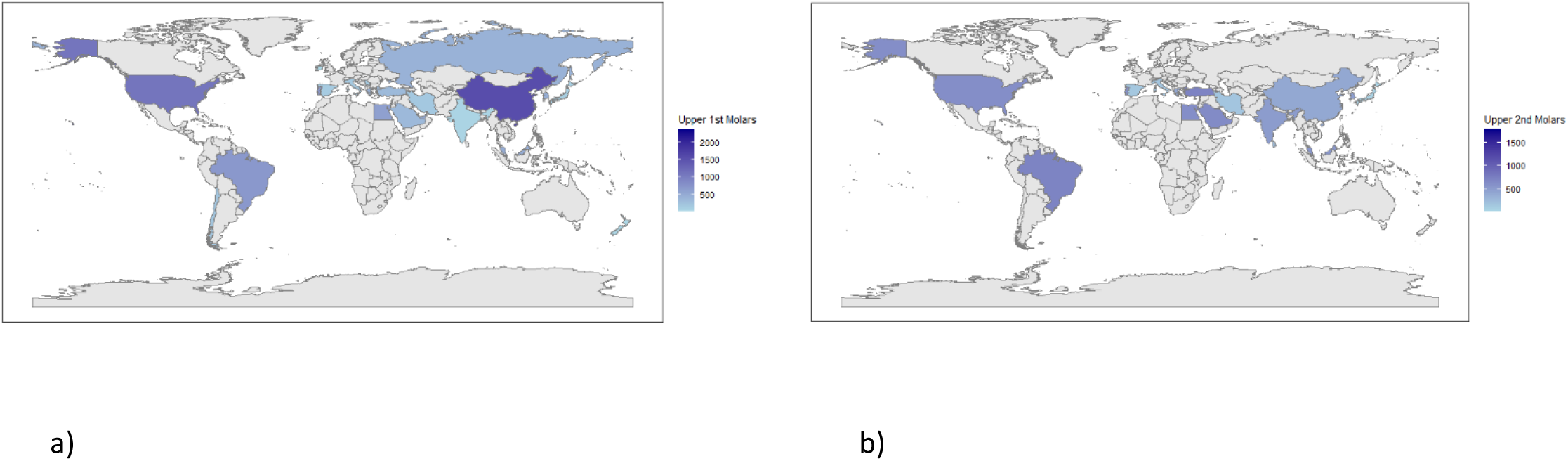
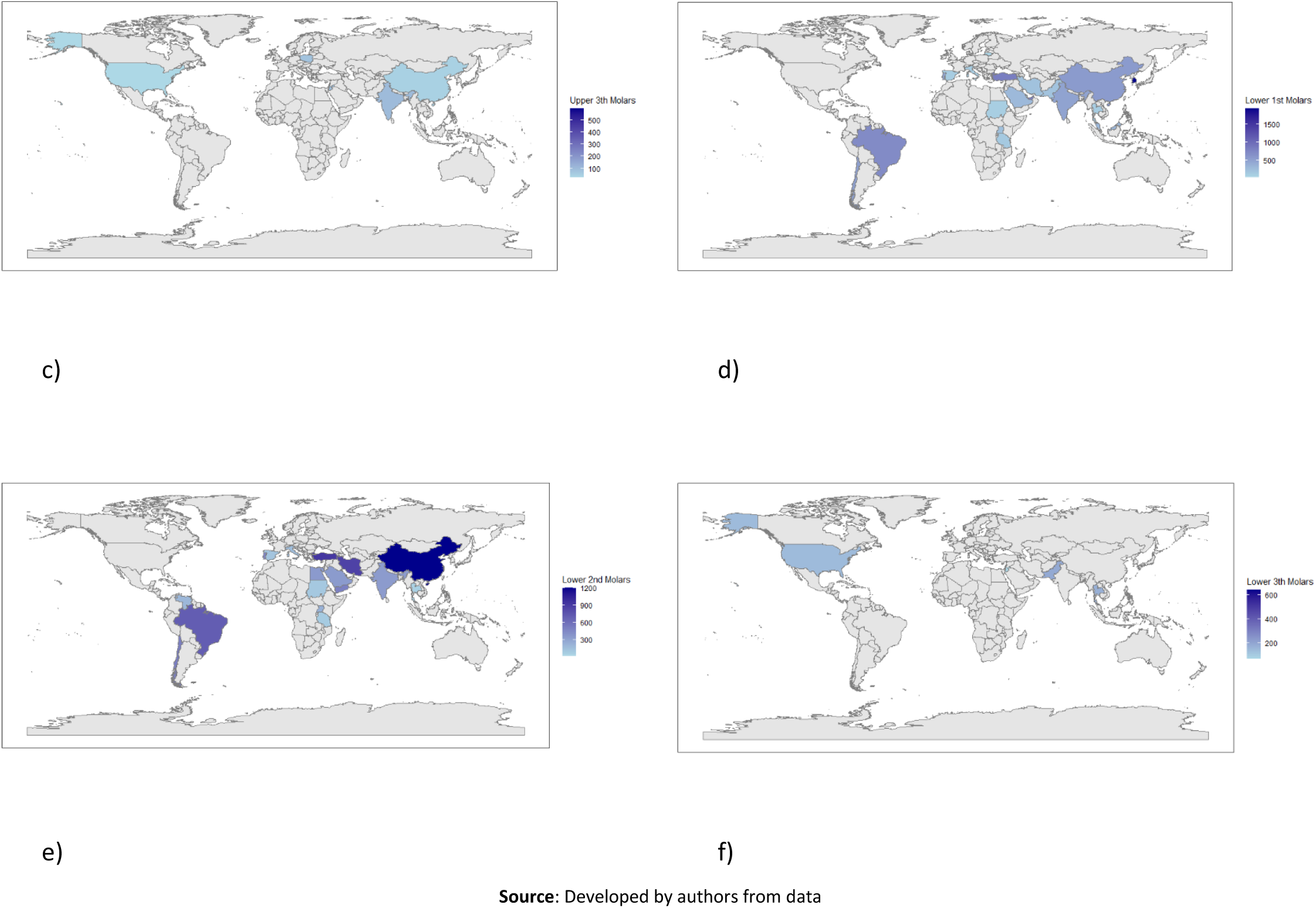
Distribution by country of the different molar groups studied. a) upper 1^st^ molar, b) upper 2^nd^ molars, c) upper 3^th^ molars, d) lower 1^st^ molars, e) lower 2^nd^ molars and f) lower 3^th^ molars.

Among the upper first molars, China was the country with the most reported molars (30.21 %) and Ireland (0.01 %), being the only dental group present in the data extracted from the studies included in the research (Table 2 and Figure 4).

In relation to the upper second molars (28.58%), China contributed 30.01% of the total collected in the studies, and Japan (0.31%) had the lowest amount. (Table 2 & Figure 4)

For the upper third molars, the only countries that contributed data in the included studies were Jordan (63.64%), the United States (15.70%), India (9.35%), Poland (7.29%), and China (4.02%). (Table 2 & Figure 4)

The lower first molar had 17.49% of the total data analyzed in the studies included in this research. South Korea had the highest number of lower first molars (18.36 %), followed by Lithuania (0.59 %). (Table 2 & Figure 4)

The lower second molar accounted for 16.96% of the total number of patients included. The countries with the most molars in this dental group were China (18.96 %) and Thailand (0.59 %). (Table 2 & Figure 4)

In the upper third molar, the only countries that contributed data in the studies included in the research were Jordan (58.16%), Pakistan (15.83%), Thailand (14.19%) and the United States (11.81%). (Table 2 & Figure 4)

According to the data obtained in the studies in relation to the number of root canals in permanent molars, it is evident that in the African continent, it is common to find the first and second upper molars with three root canals with values of 45.62% and 41.31%, respectively. The same occurred in the first and second lower molars, with values of 59.79% for the first and 67.55% for the second. According to the information from the studies included in the research, there were no reports of treatment of the upper and lower third molars on this continent. (Table 3)

**Table 3.** Distribution of the number of root canals for each molar group in relation to the continents.

|  | 1st Upper Molar |  |  |  |  |  |  |  |  |  |  |  |  |  |
| --- | --- | --- | --- | --- | --- | --- | --- | --- | --- | --- | --- | --- | --- | --- |
|  | Total of Molars |  | 1 Canal |  | 2 Canals |  | 3 Canals |  | 4 Canals |  | 5 Canals |  | 6 Canal |  |
|  | # | % | # | % | # | % | # | % | # | % | # | % | # | % |
| Africa | 605 | 3,02 | 0 | - | 0 | - | 276 | 45,62 | 175 | 28,93 | 154 | 25,45 | 0 | - |
| Americas | 3117 | 15,55 | 0 | - | 21 | 0,67 | 1314 | 42,16 | 1766 | 56,66 | 16 | 0,51 | 0 | - |
| Asia | 14159 | 70,65 | 3 | 0,02 | 124 | 0,88 | 7743 | 54,69 | 6182 | 43,66 | 97 | 0,69 | 10 | 0,07 |
| Europe | 2140 | 10,68 | 16 | 0,75 | 41 | 1,92 | 1323 | 61,82 | 732 | 34,21 | 28 | 1,31 | 0 | - |
| Oceania | 20 | 0,10 | 0 | - | 0 | - | 3 | 15,00 | 4 | 20,00 | 0 | - | 13 | 65,00 |
| Sub- Total | 20041 | 33,18* | 19 | 0,09 | 186 | 0,93 | 10659 | 53,19 | 8859 | 44,20 | 295 | 1,47 | 23 | 0,11 |
|  | 2nd Upper Molar |  |  |  |  |  |  |  |  |  |  |  |  |  |
|  | Total of Molars |  | 1 Canal |  | 2 Canals |  | 3 Canals |  | 4 Canals |  | 5 Canals |  | 6 Canal |  |
|  | # | % | # | % | # | % | # | % | # | % | # | % | # | % |
| Africa | 610 | 3,53 | 0 | - | 0 | - | 252 | 41,31 | 133 | 21,80 | 225 | 36,89 | 0 | - |
| Americas | 1649 | 9,55 | 8 | 0,49 | 60 | 3,64 | 876 | 53,12 | 703 | 42,63 | 2 | 0,12 | 0 | - |
| Asia | 13539 | 78,43 | 148 | 1,09 | 2494 | 18,42 | 7885 | 58,24 | 2970 | 21,94 | 42 | 0,31 | 0 | - |
| Europe | 1465 | 8,49 | 21 | 1,43 | 355 | 24,23 | 809 | 55,22 | 268 | 18,29 | 12 | 0,82 | 0 | - |
| Oceania | 0 | - | 0 | - | 0 | - | 0 | - | 0 | - | 0 | - | 0 | - |
| Sub- Total | 17263 | 28,58* | 177 | 1,03 | 2909 | 16,85 | 9822 | 56,90 | 4074 | 23,60 | 281 | 1,63 | 0 | - |
| **3th Upper Molar** |  |  |  |  |  |  |  |  |  |  |  |  |  |  |
| Africa | 0 | - | 0 | - | 0 | - | 0 | - | 0 | - | 0 | - | 0 | - |
| Americas | 168 | 15,70 | 4 | 2,38 | 10 | 5,95 | 101 | 60,12 | 46 | 27,38 | 6 | 3,57 | 1 | 0,60 |
| Asia | 824 | 77,01 | 74 | 8,98 | 112 | 13,59 | 421 | 51,09 | 203 | 24,64 | 14 | 1,70 | 0 | - |
| Europe | 78 | 7,29 | 18 | 23,08 | 12 | 15,38 | 42 | 53,85 | 6 | 7,69 | 0 | - | 0 | - |
| Oceania | 0 | - | 0 | - | 0 | - | 0 | - | 0 | - | 0 | - | 0 | - |
| Sub- Total | 1070 | 1,77* | 96 | 8,97 | 134 | 12,52 | 564 | 52,71 | 255 | 23,83 | 20 | 1,87 | 1 | 0,09 |
| **1st Lower Molar** |  |  |  |  |  |  |  |  |  |  |  |  |  |  |
| Africa | 470 | 4,45 | 0 | - | 94 | 20,00 | 281 | 59,79 | 92 | 19,57 | 3 | 0,64 | 0 | - |
| Americas | 1667 | 15,78 | 0 | - | 118 | 7,08 | 1002 | 60,11 | 526 | 31,55 | 18 | 1,08 | 3 | 0,18 |
| Asia | 7515 | 71,15 | 6 | 0,08 | 518 | 6,89 | 4693 | 62,45 | 2223 | 29,58 | 72 | 0,96 | 3 | 0,04 |
| Europe | 910 | 8,62 | 0 | - | 68 | 7,47 | 739 | 81,21 | 103 | 11,32 | 0 | - | 0 | - |
| Oceania | 0 | - | 0 | - | 0 | - | 0 | - | 0 | - | 0 | - | 0 | - |
| Sub- Total | 10562 | 17,49* | 6 | 0,06 | 798 | 7,56 | 6715 | 63,58 | 2944 | 27,87 | 93 | 0,88 | 6 | 0,06 |
| **2nd Lower Molar** |  |  |  |  |  |  |  |  |  |  |  |  |  |  |
| Africa | 758 | 7,40 | 11 | 1,45 | 147 | 19,39 | 512 | 67,55 | 78 | 10,29 | 10 | 1,32 | 0 | - |
| Americas | 1417 | 13,83 | 10 | 0,71 | 301 | 21,24 | 1049 | 74,03 | 57 | 4,02 | 0 | - | 0 | - |
| Asia | 7155 | 69,83 | 309 | 4,32 | 1833 | 25,62 | 4692 | 65,58 | 316 | 4,42 | 5 | 0,07 | 0 | - |
| Europe | 917 | 8,95 | 9 | 0,98 | 366 | 39,91 | 517 | 56,38 | 24 | 2,62 | 1 | 0,11 | 0 | - |
| Oceania | 0 | - | 0 | - | 0 | - | 0 | - | 0 | - | 0 | - | 0 | - |
| Sub- Total | 10247 | 16,96* | 339 | 3,31 | 2647 | 25,83 | 6770 | 66,07 | 475 | 4,64 | 16 | 0,16 | 0 | - |
| **3th Lower Molar** |  |  |  |  |  |  |  |  |  |  |  |  |  |  |
| Africa | 0 | - | 0 | - | 0 | - | 0 | - | 0 | - | 0 | - | 0 | - |
| Americas | 144 | 11,81 | 5 | 3,47 | 25 | 17,36 | 83 | 57,64 | 25 | 17,36 | 5 | 3,47 | 1 | 0,69 |
| Asia | 1075 | 88,19 | 36 | 3,35 | 461 | 42,88 | 474 | 44,09 | 103 | 9,58 | 1 | 0,09 | 0 | - |
| Europe | 0 | - | 0 | - | 0 | - | 0 | - | 0 | - | 0 | - | 0 | - |
| Oceania | 0 | - | 0 | - | 0 | - | 0 | - | 0 | - | 0 | - | 0 | - |
| Sub- Total | 1219 | 2,02* | 41 | 3,36 | 486 | 39,87 | 557 | 45,69 | 128 | 10,50 | 6 | 0,49 | 1 | 0,08 |
| **Total** | **60402** | **100** | **678** | **1,12** | **7160** | **11,85** | **35087** | **58,09** | **16735** | **27,71** | **711** | **1,18** | **31** | **0,05** |

\*These percentages are derived from the total molars at the end of the table (n=60402). The remaining percentages were at the rate of the subtotal for each dental group.
Font: Developed by authors from data

In America, there are many upper first molars with four root canals, representing 56.66% of the total molars in this dental group. The remaining dental groups had three root canals, with values of 53.12% for the second upper molar, 60.12% for the third upper molar, 60.11% for the first lower molar, 67.55% for the second lower molar, and 57.64% for the third lower molar. (Table 3)

On the Asian continent, all the dental groups included in the study had three root canals, with values of 54.69% for the first upper molar, 58.24% for the second upper molar, 51.09% for the upper third molar, 62.45% for the lower first molar, and 65.58% for the lower second molar. In the case of lower third molars, there was a predominance of three root canals, although there was a very similar percentage of lower third molars with two root canals, representing 42.88% of the total. (Table 3)

On the European continent, all dental groups were represented by permanent molars with three root canals, with the exception of the lower third molars, for which there are no reports on the number of root canals in this dental group for this continent. The values were 61.82%, 55.22 %, 53.85 %, 81.21 %, and 56.38% for the first, second, third, first, and second molars, respectively. (Table 3)

In Oceania, according to the data extracted from the studies included in the research, only reports indicate that it is more common to find upper first molars with six root canals, with a value of 65% of the total. (Table 3)

## DISCUSSION

Study sensitivity analysis is a procedure that helps researchers gauge the effectiveness of the results and guide readers on the accuracy of the information collected. In a study by *Franka Marušić et al*^(103)^ they report that only 52% of the articles reported on quality thresholds for sensitivity analysis. In contrast, *Ho et al.*^(104)^ reported that only 0.9% of the articles evaluated had high methodological quality. These findings emphasize the importance of conducting rigorous evaluations of the quality of systematic review reports to ensure the reliability of the extracted data. In our study, a small group of included articles received a low sensitivity and accuracy evaluation, corresponding to articles published at a time when high-fidelity imaging equipment such as tomographs did not yet exist. Thus, the present research has guaranteed that the findings of the included studies provide reliable results.

Ethnicity reveals significant anatomical variations in the teeth, as evidenced by the findings of several investigations. There is relevant variability in the number of root and root canal configurations dating from different historical periods, suggesting that genetics influences morphological diversity ^(7, 105)^ In addition, odontometric analyses have shown that East and Southeast Asians share dental similarities with sub-Saharan Africans, which places them at the center of a wide range of dental characteristics^(106)^ On the other hand, research on African-American populations has shown the impact of mixing, derivation, and localized gene flow on dental morphology, which demonstrates the complex relationship of sociohistorical factors when studying anatomical characteristics^(107)^ All these studies highlight the intricate correspondence between ethnic origins and dental anatomical variations and shed light on the diverse patterns observed in different population groups around the world^(7, 108)^. This background derives the complexity of establishing anatomical patterns by considering populations of different continents, although a general view is helpful in visualizing an overview of these times.

Latin Americans show intermediate frequencies of dental features compared to those of Native Americans, Europeans and Africans^(109)^ This is due to the mass exodus from the European and African continents centuries ago, which evidently had a direct impact on the anatomical characteristics of today’s inhabitants. In the present study, the American population had divergent characteristics with the European and African populations^(110)^ in terms of the maxillary first molar, but homogeneous samples were not recovered from these last two continents, as in the case of the Asian continent. The rest of the molar groups had similar typologies, although the study by *Delgado et al*^(109)^ shows that dental characteristics provide a low predictive power of individual genetic ancestry. These findings underscore the importance of taking ethnicity into account when assessing anatomical variations in the permanent molar canals.

Naseri et al.^(111)^ analyzed studies from Iran and stated that although most maxillary first molars have three roots, the studies included in this research demonstrated a high frequency of two or more ducts in the mesiobuccal root. In the current study, the results concurred in a similar manner, and although the first maxillary molars with three canals prevailed, the second highest prevalence found in the studies was that of molars with four root canals. These results indicate that the variability between three and four canals in clinical practice can be frequently found according to geographical context or globally. This was demonstrated by a systematic review by Barbhai et al.^(112)^, who found that 68.2% of the first maxillary molars had a second mesiobuccal canal.

A systematic review by *Valencia de Pablo et al.*^(113)^ attempted to analyze the published literature related to root anatomy and configuration of the root canal system of the mandibular permanent first molar. In relation to the number of canals in this dental group, they determined that of 18 studies that included 4745 teeth, 61.3% had three canals, 35.7% 4 canals and about 1% had five canals. These results are almost identical to those obtained in the present study, and it is worth noting that they included reports with the same geographical diversity as this study.

*Valencia de Pablo et al*^(113)^ suggest that the number of roots of the mandibular first molar is directly related to the ethnicity of the population studied. The presence of a third root is more common in the Native American, Eskimos, and Chinese populations, suggesting a genetic component.

Another systematic review by *AL-Rammahi et al.(*^(114)^ observed the presence of a medial mesial canal in some studies, with a variable prevalence, which implies that the alterations of the normal anatomy of the mandibular first molar conform more to an anatomical pattern of three canals, as classically expressed in the academic literature. In addition, in distal roots, the study found a conduit that corresponded to the most common configuration. These results coincide with the prevalence of mandibular first molars in the three canals in the present study, although it is essential to take into account the anatomical variability of this molar group to obtain successful endodontic treatment.

Although the research by *Mashyakhy et al.*^(115)^ was exclusive to studies that included mandibular teeth from Saudi Arabian populations, it was found that the first mandibular molars had three and four canals with a higher prevalence (58.7% and 40.6%, respectively). In the present study, the results were convergent with those of *Mashyakhy et al.*^(115)^ even when these characteristics were specified in the continent to which that nation belongs.

In the study by Joshi et al.^(116)^, the mandibular second molars had three roots. In their systematic review, most mesial roots had two canals (70.4%) and most distal roots had a single canal (77%). *Mashyakhy et al.*^(115)^ found similar results in their systematic review research exclusively for population studies in Saudi Arabia. The present research had similar global and individual results for each continent on the prevalence of mandibular second molars with three canals, although in all the studies that studied this molar group there were cases of teeth that had two canals with a not so low prevalence. This fact warns that the anatomy of the root canals in the second molars of the mandible is variable and complex.

Limited research on the root and root canal morphology of third molars faces important implications for dental practice and the understanding of craniofacial morphology. Studies have shown that third molars have an unpredictable anatomy, with new canal configurations that had not been previously described in the literature, which highlights the need for further research^(4)^ In addition, craniofacial morphology has been related to the impaction of the third molars^(117)^ and their agenesis^(118)^ which exposes the importance of taking into account other facial parameters in treatment decisions.

The study by *Morita et al*^(119)^ encourages understanding the metameric variation in human molars through a detailed morphological analysis that can provide information on the evolution and development of teeth. Perhaps applied to this dental group it can allow a better understanding of its dental morphology and its evolutionary implications.

The findings of this study suggest that the internal anatomy of permanent molars, particularly the number of root canals, presents significant variations globally. These variations have important implications in clinical practice, as they can influence endodontic treatment planning, the likelihood of treatment success, and the management of complications. More research is needed to understand the causes of these variations and develop more precise treatment strategies for different populations.

Among the limitations of the present study is the fact that although all continents were representative and the sample was large enough to generalize the results, there was no homogeneity in the distribution due to lack of studies in many of the continents. Such was the case in America, specifically North America, Africa and Europe. Despite this, it was possible to visualize a pattern in the number of canals for each molar group studied and to delimit them to the geographical regions in question.

The future directions for this controversial topic in the dental profession aim to systematize in the compilation of new inclusive studies for a subsequent analysis of the anatomical behavior of the root canals of posterior teeth and to the need to carry out studies to make a sketch of what would be interesting from the point of view of the clinical view focused in the presence of number of root canals have those teeth when the clinician performs approaches for endodontic treatments.

## CONCLUSIONS

The study evaluated geographic variation in the internal anatomy of the root canals of permanent molars and found a wide variability in the number of root canals according to geographic region. The results reveal distinct patterns in the Americas, Asia, Africa, Europe, and Oceania, with varying prevalences of different root canal configurations on each continent.

It was observed that, in Africa, the upper and lower first and second molars most frequently have three root canals. In America, the first upper molars stand out for having four root canals. In Asia, most molars have three root canals, except for the lower third molars, where a similar proportion of two and three root canals is observed. In Europe, the upper and lower molars, except for the lower third, show a predominance of three root canals. Finally, in Oceania, the first upper molars are characterized by having six root canals with a higher frequency.

## Data Availability

All data produced are available online at Mendely Data.

https://doi.org/10.17632/xvgf59n75k.1

## Conflict of interest

The authors declare that they have no conflicts of interest.

## Funding

This research was developed using the researchers’ own resources and did not receive institutional funding.

## AUTHORSHIP CONTRIBUTION

**Concepts**: Alain Manuel Chaple Gil.

**Data curation**: Alain Manuel Chaple Gil, Meylin Santiesteban Velázquez.

**Analysis**: Alain Manuel Chaple Gil, Meylin Santiesteban Velázquez, Kelvin Ian Afrashtehfar.

**Research**: Alain Manuel Chaple Gil, Meylin Santiesteban Velázquez, Kelvin Ian Afrashtehfar.

**Methodology**: Alain Manuel Chaple Gil

**Software**: Alain Manuel Chaple Gil

**Supervision**: Alain Manuel Chaple Gil

**Validation**: Alain Manuel Chaple Gil, Kelvin Ian Afrashtehfar.

**Display:** Alain Manuel Chaple Gil, Meylin Santiesteban Velázquez, Kelvin Ian Afrashtehfar

**Writing - original draft**: Alain Manuel Chaple Gil, Meylin Santiesteban Velázquez, Kelvin Ian Afrashtehfar

**Drafting - revision and editing**: Alain Manuel Chaple Gil, Meylin Santiesteban Velázquez, Kelvin Ian Afrashtehfar.

---

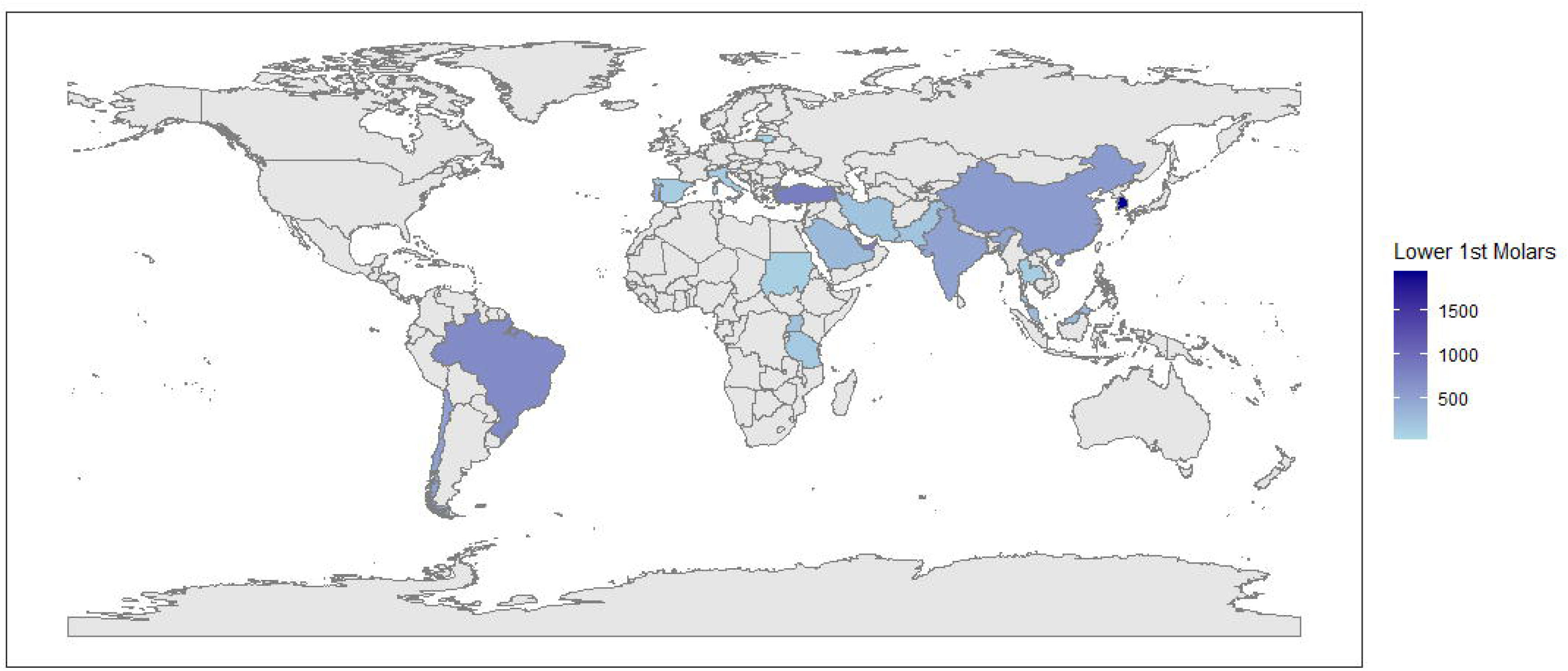
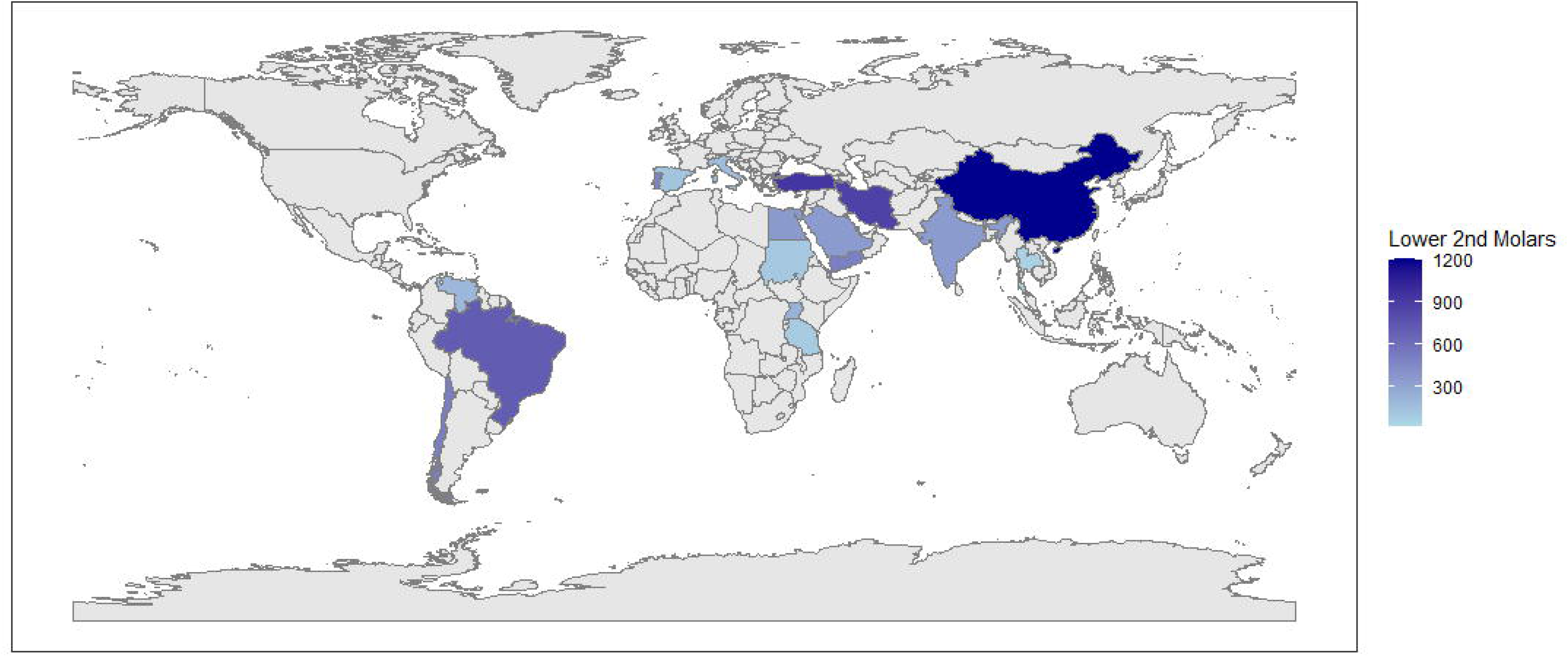
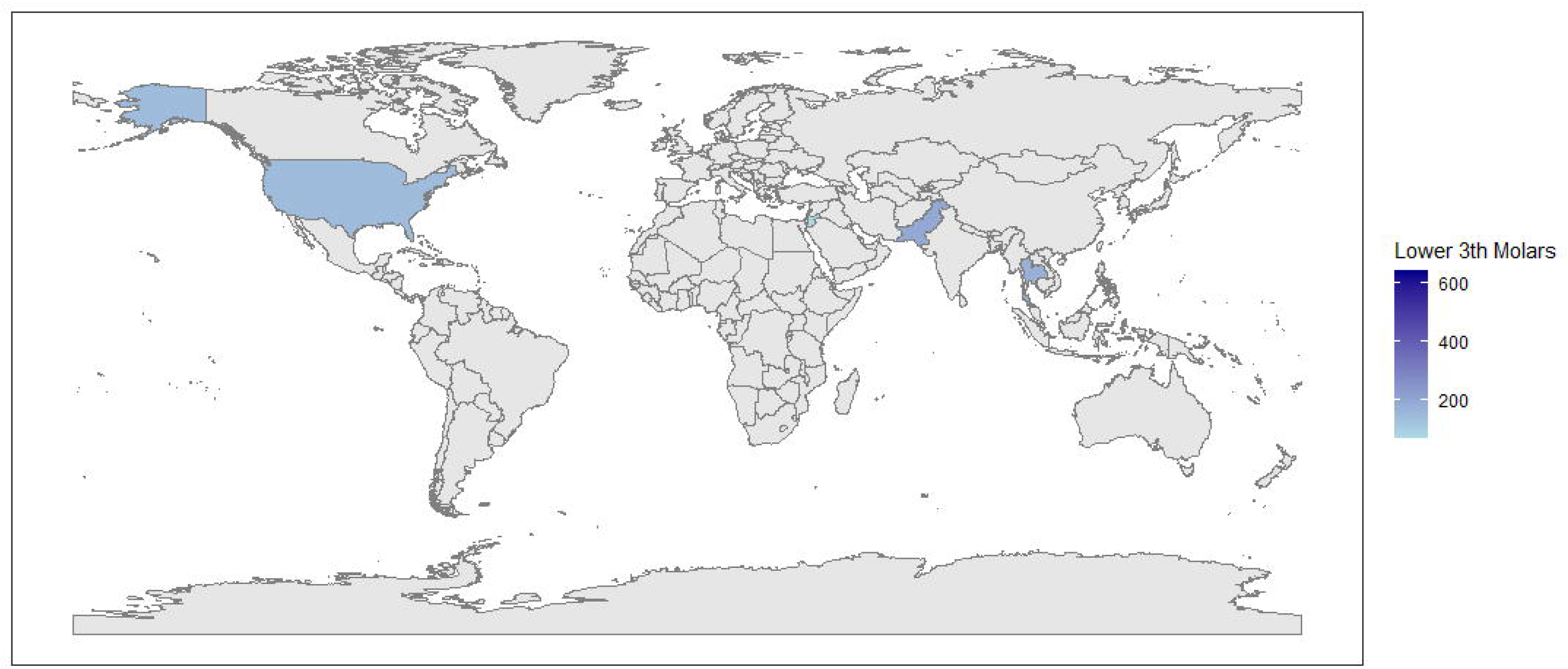
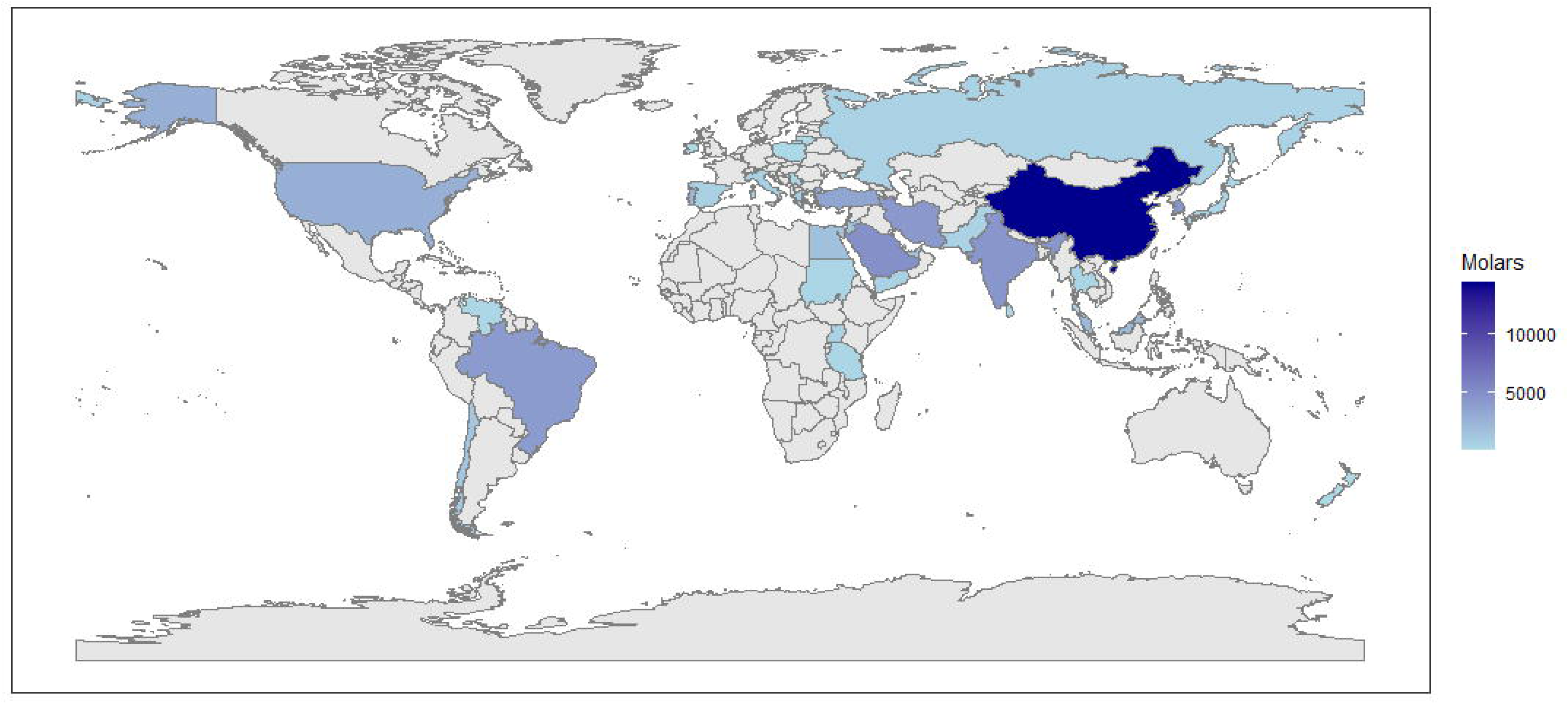
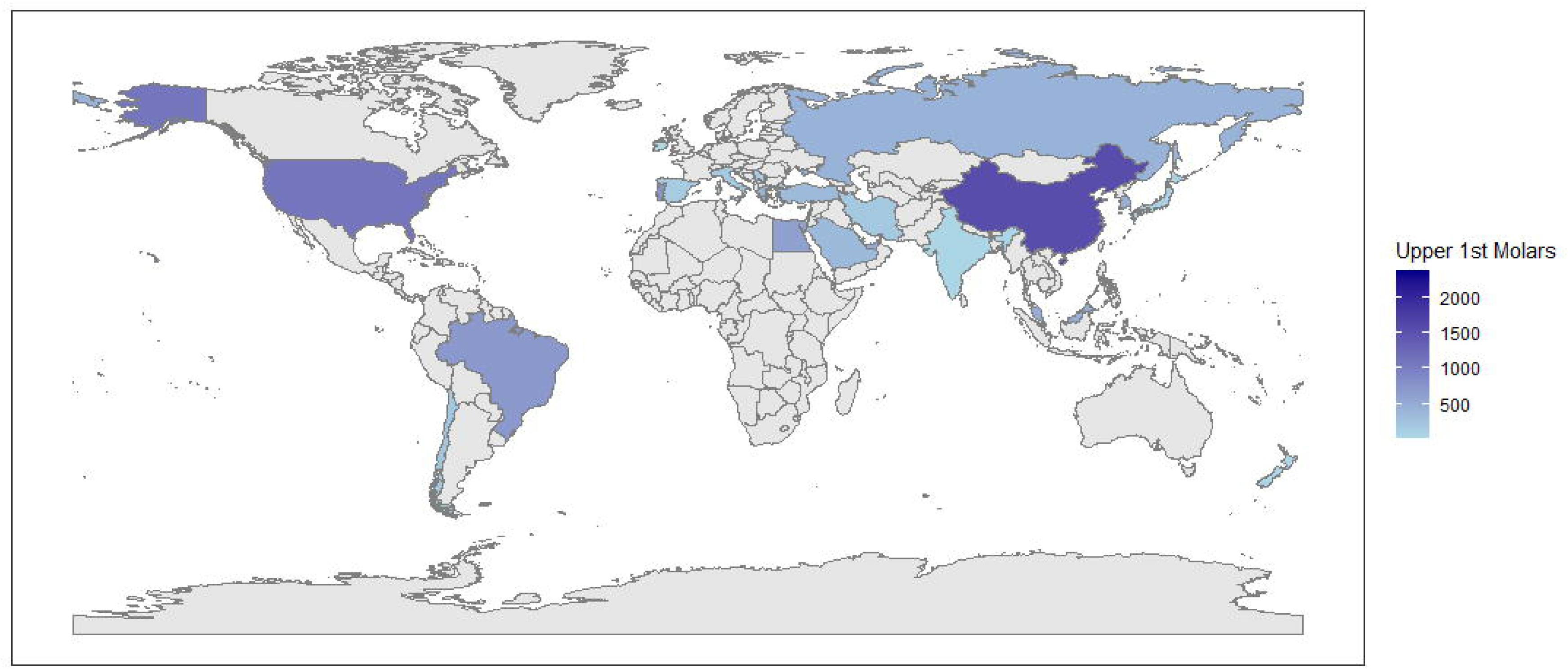
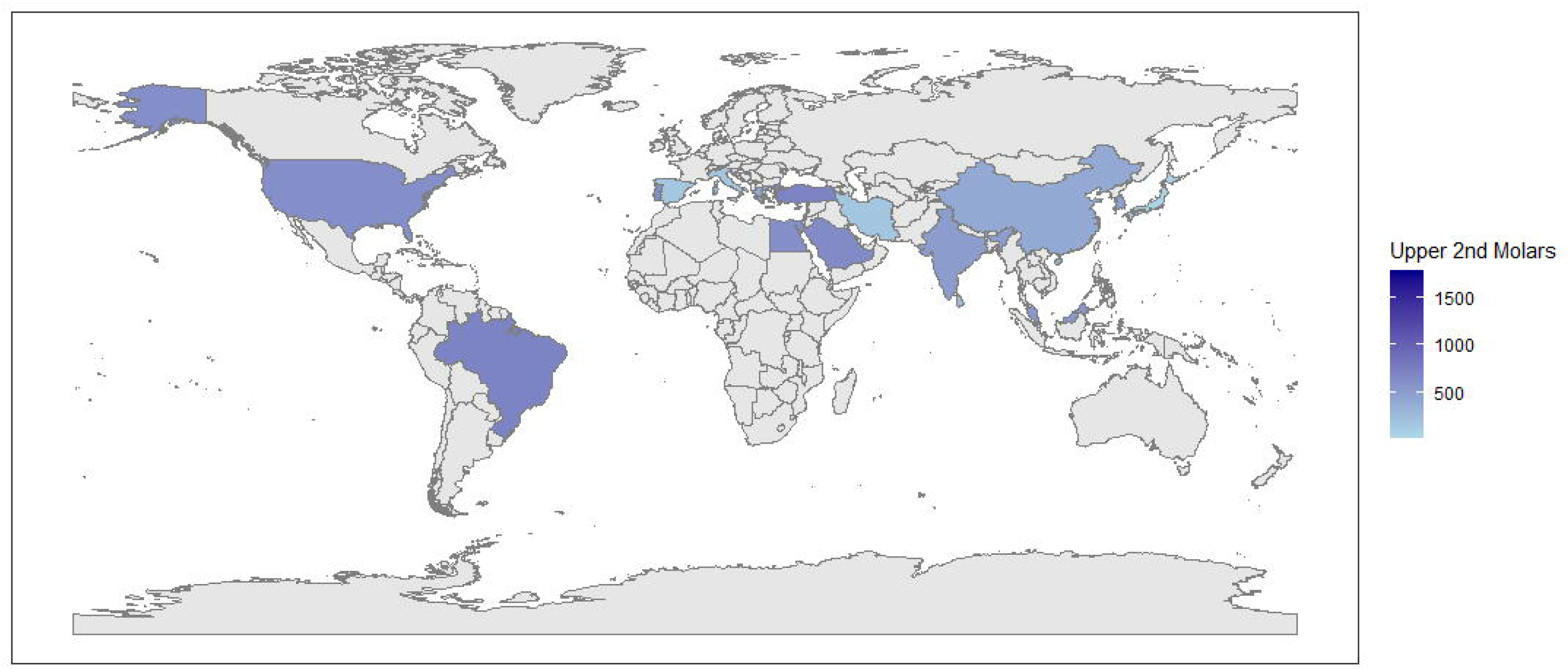
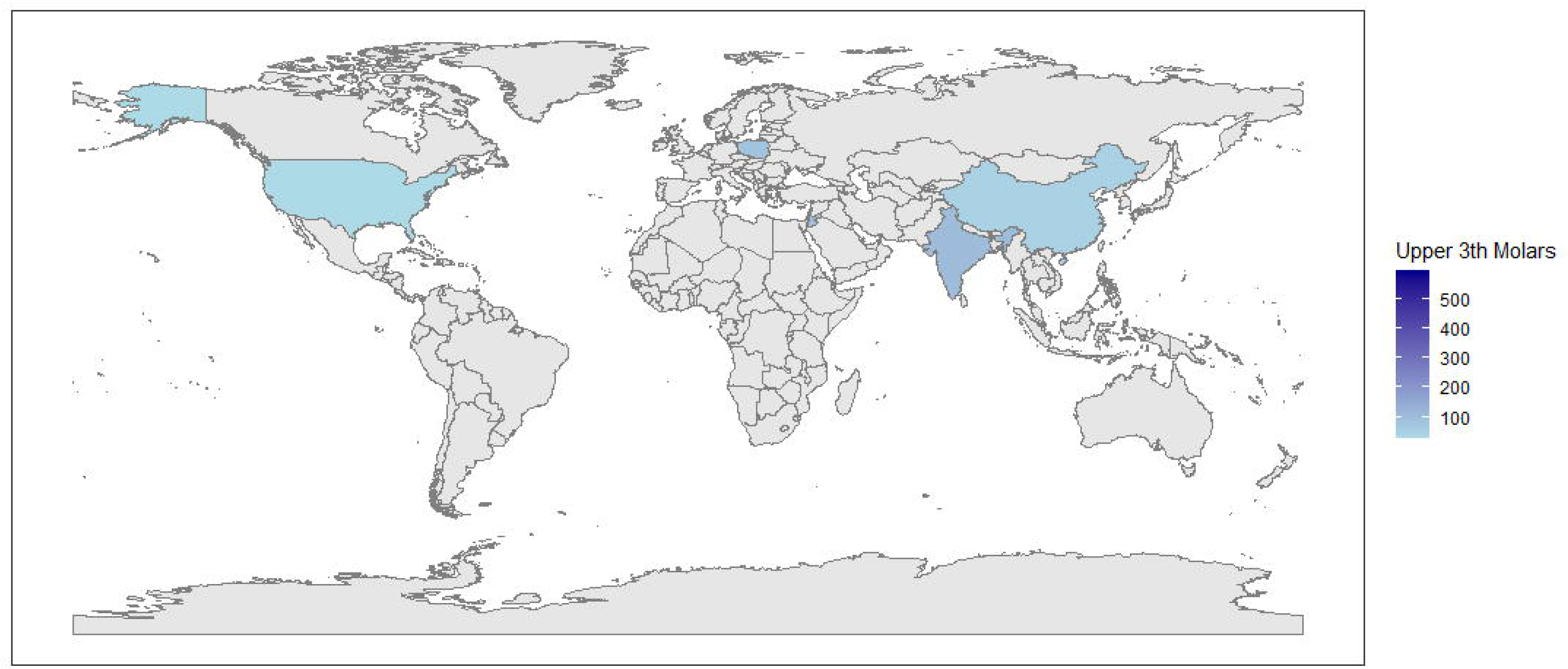

## Notes

### Competing Interest Statement

The authors have declared no competing interest.

### Funding Statement

This study did not receive any funding

